# Weather variation in objectively measured physical activity: results of four Brazilian cohorts

**DOI:** 10.1101/2023.03.02.23285008

**Authors:** Rafaela Costa Martins, Cauane Blumenberg, Andrea T Wendt, Werner de Andrade Müller, Iná S Santos, Alicia Matijasevich, Marlos Domingues, Andréa D Bertoldi, Helen Gonçalves, Fernando C. Wehrmeister, Felipe Fossati Reichert

**Affiliations:** Post-graduate program in Epidemiology, Federal University of Pelotas, Brazil; causale consultoria, Pelotas, Brazil; Post-graduate program in Epidemiology, Federal University of Pelotas, Brazil; Post-graduate Program of Physical Education, Federal University of Pelotas, Brazil; Departamento de Medicina Preventiva, Faculdade de Medicina FMUSP, Universidade de São Paulo, SP, Brasil

**Keywords:** Physical Activity, Epidemiology, Accelerometry, Motion Sensors, Exercise

## Abstract

**Background:** The aim of the present study was to examine the relationship of objectively measured physical activity and weather variables in different stages of life course.

**Methods:** This study used data from four Brazilian cohorts (mean ages: 10.9, 22.6, 30.2, and 70.5 years). The exposure was weather variation, using temperature, rain, humidity, and wind velocity. The outcome measured was accelerometer based physical activity in three ways: overall, 5-minute bouts, and 10-minute bouts physical activity. Both exposure and outcome were collected from the same period. Crude and adjusted analyses were fitted using a two-part model.

**Results:** Among 9,966 individuals from different ages evaluated, physical activity levels were low, especially in older adults. Rain and mean wind velocity were not associated physical activity of individuals, however, the probability of children being inactive was higher during humid days. After mutually adjusting the models for weather variables, only mean temperature and humidity were associated with higher and lower physical activity levels in children, respectively.

**Conclusions:** Weather variables are not key indicators to be accounted when modelling physical activity studies in almost all ages. However, this study helps to identify specifically exposures, regardless of the physical activity operationalization.

## 1. Introduction

Physical activity effects on health are well established, since it can prevent premature mortality(1) and a series of chronic medical conditions.(2) Evidence also supports the reduction of cardiometabolic diseases, different types of cancer and an improvement in mental health and quality of life.(3) Despite this, high prevalence of physical inactivity is globally observed(4) and there is no evidence of increase in physical activity over the years.(5) As physical activity is a multifactorial behavior,(6) the increase in its prevalence depends on several approaches.(7) Ecological models explored the influence of individual and environmental dimensions on lifestyle behavior, including policies, social, and natural factors.(8) It is important to notice that most studies considering physical activity rely on individual determinants, and climate characteristics are less explored.(9,10)

Weather and climate indicators (e.g., precipitation, temperature, humidity, wind velocity) can affect physical activity, acting as incentives or barriers in different populations.(11) Data from 23,451 participants in the International Children’s Accelerometry Database (ICAD), obtained from 21 worldwide studies, showed higher averages of physical activity in children and adolescents exposed to more hours of daylight, better visibility and higher temperatures, while the lower averages of physical activity was found for more exposure to precipitation and wind.(12) Among adults and older adults, higher physical activity levels were also positively associated with high temperatures and sunlight, for example, and negatively with precipitation and low temperatures.(13–17)

Controversial effects of the relationship between physical activity and weather conditions have also been observed in children and adults.(18–20) Extreme temperatures(21) and aspects such as dry or moist tropical conditions(22) can impact on physical activity levels. Brazil, which is usually considered a tropical country, has an extensive territory with five predominant climatic zones delimited by an enormous variability in terms of temperatures, precipitation and humidity.(23) In Southern Brazil, where our study was carried out, there is a notable thermal oscillation and, unlike other regions of the country, well defined seasons.(23)

In view of the importance of studies in low- and middle-income countries to increase physical activity,(6) it is relevant to examine weather predictors in areas with inadequate city planning, unsafety, and high levels of inequity. Indeed, in several studies already have identified the effect of weather in physical activity levels, but this topic is not so explored in low- and middle-income countries as Brazil where the opportunities to physical activity practice are marked by inequalities and lack of accesses to public/free facilities with good quality for the poorest population.(24,25) In addition, the use of more precise measures as accelerometers to assess the seasonal variation of physical activity has been applied mainly in countries of the Northern hemisphere and New Zealand, but not in South America.(11,26) Thus, the aim of the present study was to examine the relationship of objectively measured physical activity and weather variables in different stages of life course.

## 2. Materials and methods

The study included data from three birth cohorts established in Pelotas, Southern Brazil. In 1982, 1993 and 2004 all live-born children in hospitals of the city, and whose mothers lived in the Pelotas’ urban area, were eligible to participate. Furthermore, a cohort of older adults living in Pelotas (“COMO VAI?” study) that started in 2014 was also included in this study. Each cohort followed-up their participants in different time points, and data from the 11 year- (2004 birth cohort participants collected in 2015), 22 year- (1993 birth cohort participants collected in 2015-2016) and 30-year follow-ups (1982 birth cohort participants collected in 2012-2013), as well as data from the baseline of the “COMO VAI?” study, were used. More details about the methodology of the individual studies are available elsewhere(27– 31). Individuals from each cohort are, hereafter, referred as children when analyzing data from the 2004 birth cohort, young adults when analyzing data from the 1993 birth cohort, adults when analyzing data from the 1982 birth cohort, and older adults when analyzing data from the “COMO VAI?” study.

Objectively measured physical activity was collected through accelerometers placed in the non-dominant wrist during a 24-hour protocol lasting up to seven days. The devices measured raw acceleration expressed in gravitational equivalent units (1000 *m*g = 1 g = 9.81 m/s^2^). GENEActiv model(32) at a 85.7Hz frequency was used in “COMO VAI?” study and 1982 cohorts, while Actigraph (wGT3X-BT, wGT3X and ActiSleep models) at a 60Hz frequency was used in 1993 and 2004 birth cohorts. All devices were waterproof, triaxial (vertical, horizontal front-back and horizontal right-left axis) and present a strong correlation for the vector magnitude (>0.95)(33). According to Rowlands et al. (2018),(34) there is a good agreement between raw data from both devices if worn in the non-dominant wrist (intraclass correlation coefficient = 0.86; 95%CI: 0.56 – 0.95).

Accelerometers were set up and downloaded in the GENEActiv and ActiLife software, and accelerometer data was analyzed using the GGIR R-package (http://cran.r-project.org)(35). A 5-second epoch was set in all follow-ups, with the standard filter. The steps of signal processing scheme were as follows: 1) verification of sensor calibration error using local gravity as reference; 2) detection of sustained abnormally high values compatible with non-human movement and non-wear detection; 3) calculation of vector magnitude of activity-related acceleration using the Euclidian Norm minus 1g 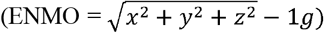 with any negative values rounded up to zero; and 4) imputation of invalid data segments by the average of similar time-of-day data points on different days of the measurement. At the end, files were considered appropriate for analyses if post-calibration error was lower than 0.02g and if there was valid data for every 15 minutes during a 24-hour cycle.

Three physical activity variables were generated: a) overall physical activity (ENMO), expressed by the daily average of acceleration in *m*g; b) moderate-to-vigorous physical activity (MVPA) with 5-minute bouts in minutes per day; and c) moderate-to-vigorous physical activity (MVPA) with 10-minute bouts in minutes per day. To be considered moderate to vigorous physical active, activities had to achieve acceleration higher than 100 mg per day similarly to previous studies.(33,36) Bouts were defined as consecutive periods in which participants spent at least 80% of this time in MVPA. Children and young adults providing fewer than three days of measurement were excluded from the analyses; for adults and older adults, participants with less than two days of measurement were excluded.

The following individual-level variables were collected during face-to-face interviews and included as independent variables: sex (male/female) and age (2004 birth cohort: 10.1 - 11.7 years; 1993 birth cohort: 21.9 - 23.5 years; 1982 birth cohort: 29.4 - 31.2 years; “COMO VAI?” study: >60 years). Also, the following city-level weather variables (mean temperature, rain, humidity, and mean wind velocity) were extracted from an official Brazilian source of weather in Pelotas (Brazilian Agricultural Research Corporation – *Embrapa*).(37) From the first day until the seventh day of accelerometer use, daily weather variables were combined as weekly means (same days of accelerometer use) for each participant. The weather exposure variables were dichotomized in <90^th^ percentile and ≥90^th^ percentile, in order to test the difference of physical activity levels in extreme weather conditions.

Descriptive analyses are presented as absolute and relative frequencies for sex, means with respective standard deviations (SD) for age and weather variables, and as medians with respective interquartile range (IQR) for physical activity variables due to its asymmetric distribution. Kruskal-Wallis test was used to compare medians of independent samples of physical activity. Crude and adjusted analyses were performed to assess the influence of weather changes on physical activity levels. For that purpose, two-part model(38) were fitted. This model is used when we have a sample with lots of 0, and it comprises two parts: logistic and linear. In the logistic part we divided the sample as “0” for the ones with 0 minutes of physical activity or “1” for the ones with >0 minutes of physical activity, indicating the probability of the individual to be inactive – this part is used only for the MVPA (5- and 10-minutes bout), since ENMO variable has a symmetric distribution. The linear part was applied only for those individuals that had >0 minutes of physical activity; the outputs were continuous variables in ENMO and MVPA (5- and 10-minutes bout). In the linear model, MVPA with 5- and 10-minute bouts were log-transformed to remove skewness, and then back-transformed to their original values to be presented as minutes per day calculated by linear combinations using the *lincom* Stata command. A significance level of 5% was considered for all analyses.

The Ethics Committee of the Faculty of Medicine of the Federal University of Pelotas, affiliated with the Brazilian National Council for Research Ethics (CONEP) of the Ministry of Health, approved all cohort follow-ups (2004 Cohort: 889.753; 1993 Cohort: 1.250.366; 1982 Cohort: 16/2012; “COMO VAI?” study: 201324538513.1.0000.5317). All individuals were informed about the goals of the study and were asked to sign a consent form. Participants that were 18 years or younger were asked to be accompanied by the legal guardian, and the responsible for that person had to sign the consent form (in the 2004 Cohort the adolescents also signed a consent form). All analyses were performed using Stata software version 15.0 (College Station, TX: StataCorp LLC).

## 3. Results

The sample size followed-up in each cohort and the number of individuals analyzed from each cohort varied, since only those with complete information for the outcome (objectively measure physical activity) were considered in the analyses: children (n followed-up = 3,565; n analyzed = 3,348), young adults (n followed-up = 3,810; n analyzed = 2,907), adults (n followed-up = 3,701; n analyzed = 2,740) and older adults (n followed-up = 1,285; n analyzed = 971). The sociodemographic characteristics of the analyzed individuals were compared to those excluded from analyses, showing differences on age for elders (data not shown; older individuals participated less than younger [p=0.001]), skin color in all four cohorts, on sex for young adults, and according to asset index for adults (Supplementary Table 1).

The individual’s sociodemographic characteristics are presented in Table 1. Most young adults, adults and older adults included in the analyses were female. In contrast, the sample of children analyzed was mostly represented by males. The mean ages and standard deviation for children, young adults, adults and older adults were, respectively, 10.9 (0.3), 22.6 (0.3), 30.2 (0.3), 70.5 (7.8) years. In Table 1, weather variables measured during the periods in which participants wore accelerometers are also presented. During their follow-up, young adults were exposed to the highest levels of rain, temperature, and maximum wind velocity compared with the other age groups. For humidity, both young and older adults experienced more humid days while using the accelerometer.

**Table 1.** Individual and weather description in Pelotas - Brazil - during the follow-up of each cohort.

|  | Children |  | Young adults |  | Adults |  | Older adults |  |
| --- | --- | --- | --- | --- | --- | --- | --- | --- |
|  | N | % | N | % | N | % | N | % |
| Sex |  |  |  |  |  |  |  |  |
| Male | 1722 | 51.4 | 1398 | 48.1 | 1329 | 48.5 | 367 | 37.8 |
| Female | 1626 | 48.6 | 1509 | 51.9 | 1411 | 51.5 | 604 | 62.2 |
| **Total** | **3,348** |  | **2,907** |  | **2,740** |  | **971** |  |
|  | Mean | SD | Mean | SD | Mean | SD | Mean | SD |
| Age (in years) | 10.9 | 0.3 | 22.6 | 0.3 | 30.2 | 0.3 | 70.5 | 7.8 |
| Mean temperature (°C) |  |  |  |  |  |  |  |  |
| <90th percentile (cooler) | 17.4 | 4.0 | 19.1 | 3.9 | 15.7 | 4.3 | 16.1 | 3.2 |
| ≥90th percentile (warmer) | 24.7 | 0.6 | 25.6 | 0.9 | 24.7 | 1.2 | 24.5 | 1.2 |
| Rain (mm) |  |  |  |  |  |  |  |  |
| <90th percentile (less rain) | 1.0 | 2.3 | 3.7 | 7.5 | 1.2 | 2.8 | 1.1 | 2.4 |
| ≥90th percentile (more rain) | 44.0 | 25.1 | 54.2 | 16.9 | 26.8 | 17.7 | 27.7 | 12.4 |
| Humidity (%) |  |  |  |  |  |  |  |  |
| <90th percentile (less humid) | 82.4 | 7.6 | 84.8 | 7.6 | 79.5 | 8.2 | 84.1 | 6.9 |
| ≥90th percentile (more humid) | 95.3 | 1.7 | 96.3 | 1.0 | 94.0 | 2.0 | 96.3 | 1.1 |
| Mean wind velocity (km/h) |  |  |  |  |  |  |  |  |
| <90th percentile (less wind) | 6.0 | 2.4 | 6.5 | 2.6 | 2.5 | 1.4 | 3.9 | 2.1 |
| ≥90th percentile (more wind) | 13.0 | 1.4 | 14.8 | 1.5 | 6.9 | 2.1 | 12.4 | 2.9 |

Figures 1A, 1B and 1C present, respectively, the ENMO levels, MVPA with 5-minutes bout, and MVPA with 10-minutes bout for each cohort. Significant differences (p<0.001) on physical activity levels were found between the different ages for all physical activity measures: ENMO, MVPA 5-minute bout, and MVPA 10-minute bout. Children’s physical activity levels were the highest for all physical activity measures, while older adults had the lowest physical activity levels.

**Figure 1.**
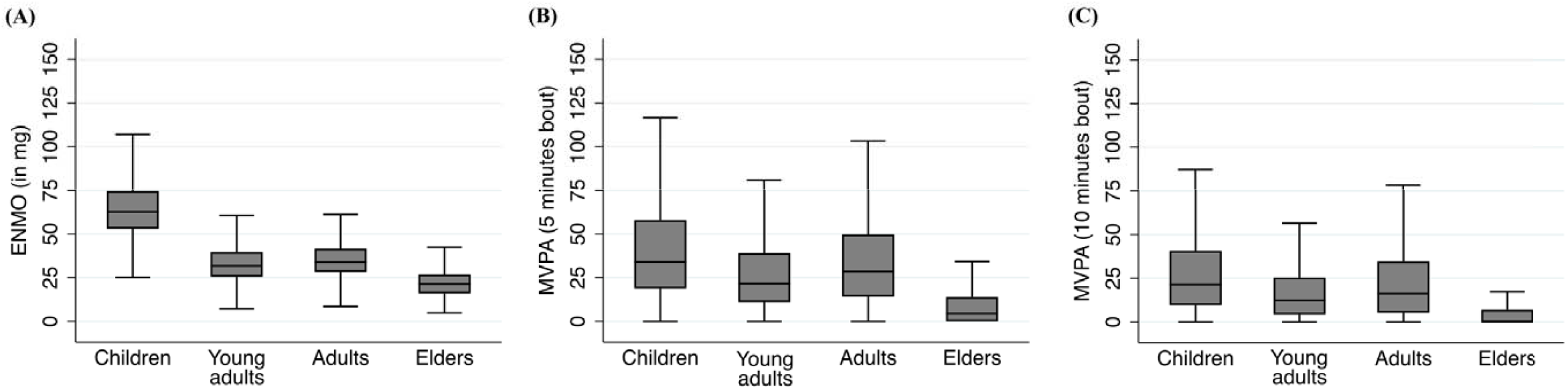
Median and interquartile intervals of physical activity by age – (A) ENMO, (B) MVPA-5 and (C) MVPA-10.

In crude analysis (Table 2), the probability of being inactive were not associated with the temperature, rain, humidity, or wind velocity regardless of the cohort or physical activity measure being analyzed. The only exception was for MVPA 10-minutes bout among children, which showed that the probability of being inactive was 1.9 times higher (95%CI = 1.2 – 3.0) in more humid than in less humid days. Linear regression showed that children that wore accelerometers during data collection spent, on average, 3.6 less minutes in more humid days than in less humid days, considering MVPA 10-minutes bout, compared to their counterparts. There was a positive association between children’s ENMO levels and mean temperature and a negative association for rain and humidity. A similar pattern was found when analyzing MVPA (5- and 10-minutes bout).

**Table 2.**
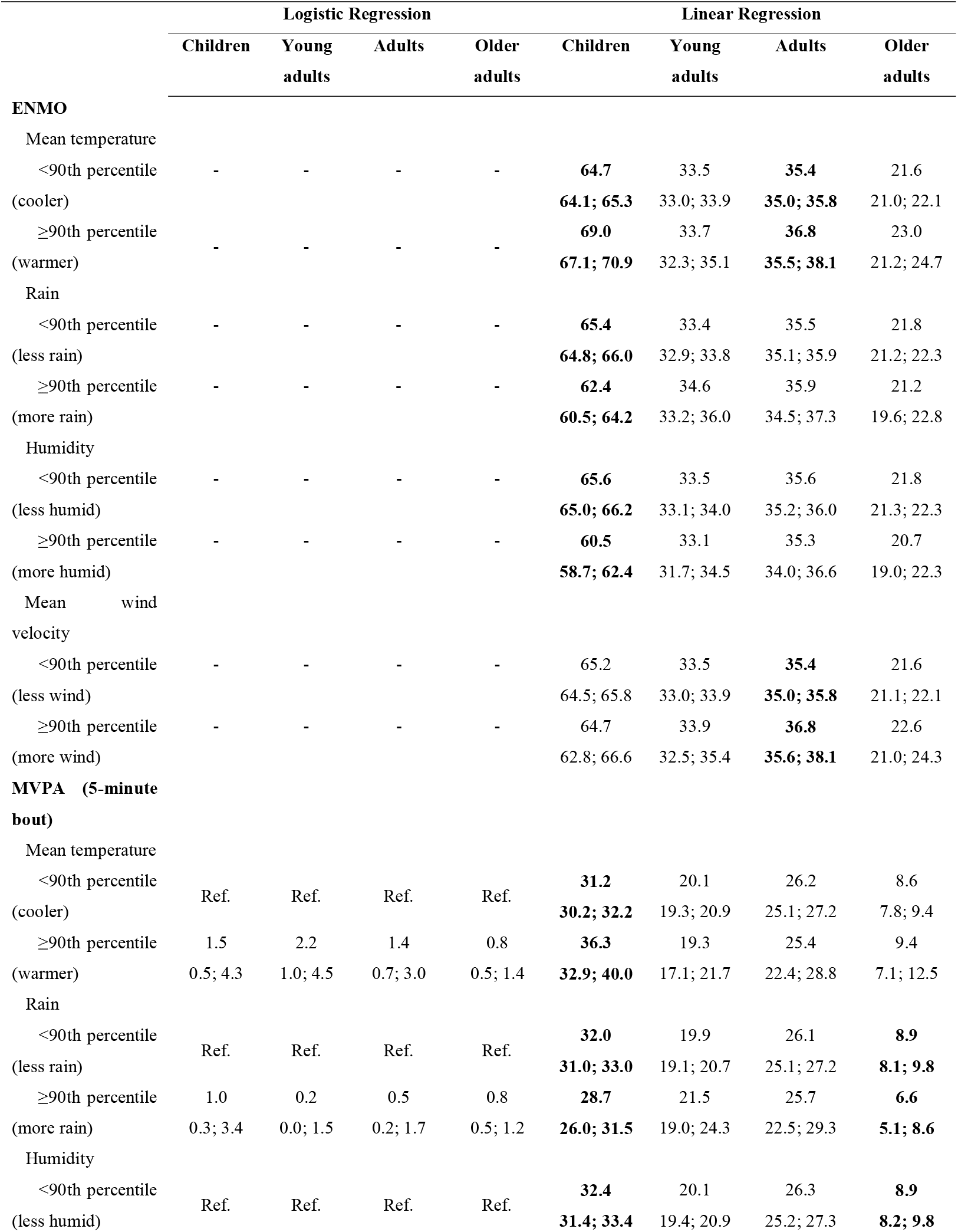
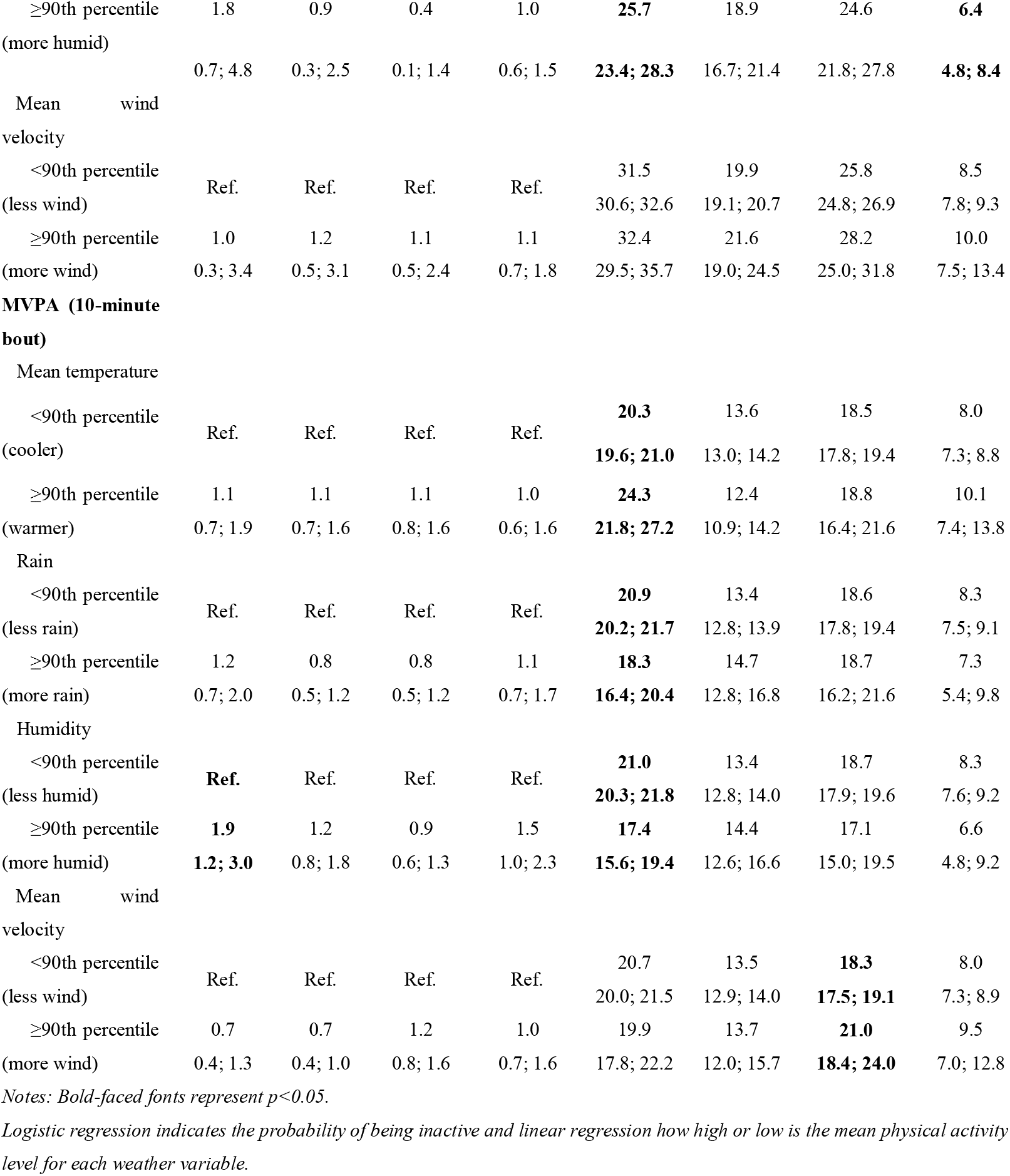
Crude association between weather variables and physical activity according to age estimated by two-part model.

Among adults, ENMO was higher in warmer and more windy days. Whereas for MVPA 5-minute bout no associations were found, and for MVPA 10-minute bouts the only association found was a direct relationship with mean wind velocity. In contrast, time spent on MVPA 5-minute bouts physical activity by older adults were significantly lower during days of extremely higher rain and humidity (Table 2).

After adjustment for weather variables, the probability of being inactive did not change – remaining significant only for MVPA 10-minute bouts among children (Table 3). In contrast, weather variables remained associated to the time spent on physical activity only among children. Physical activity was higher in warmer days and lower in more humid days for all three physical activity measures (ENMO, MVPA 5- and 10-minute bouts).

**Table 3.**
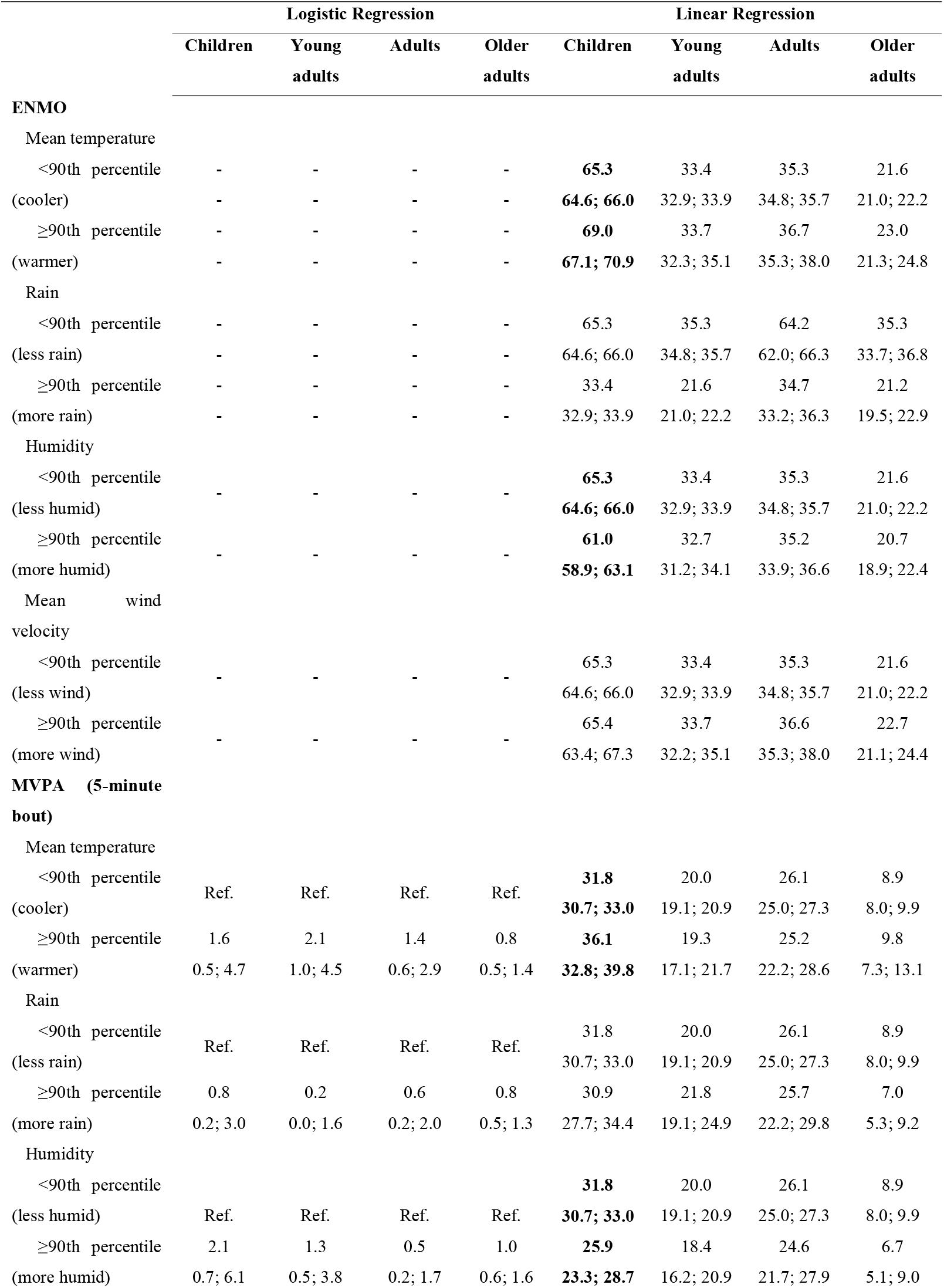
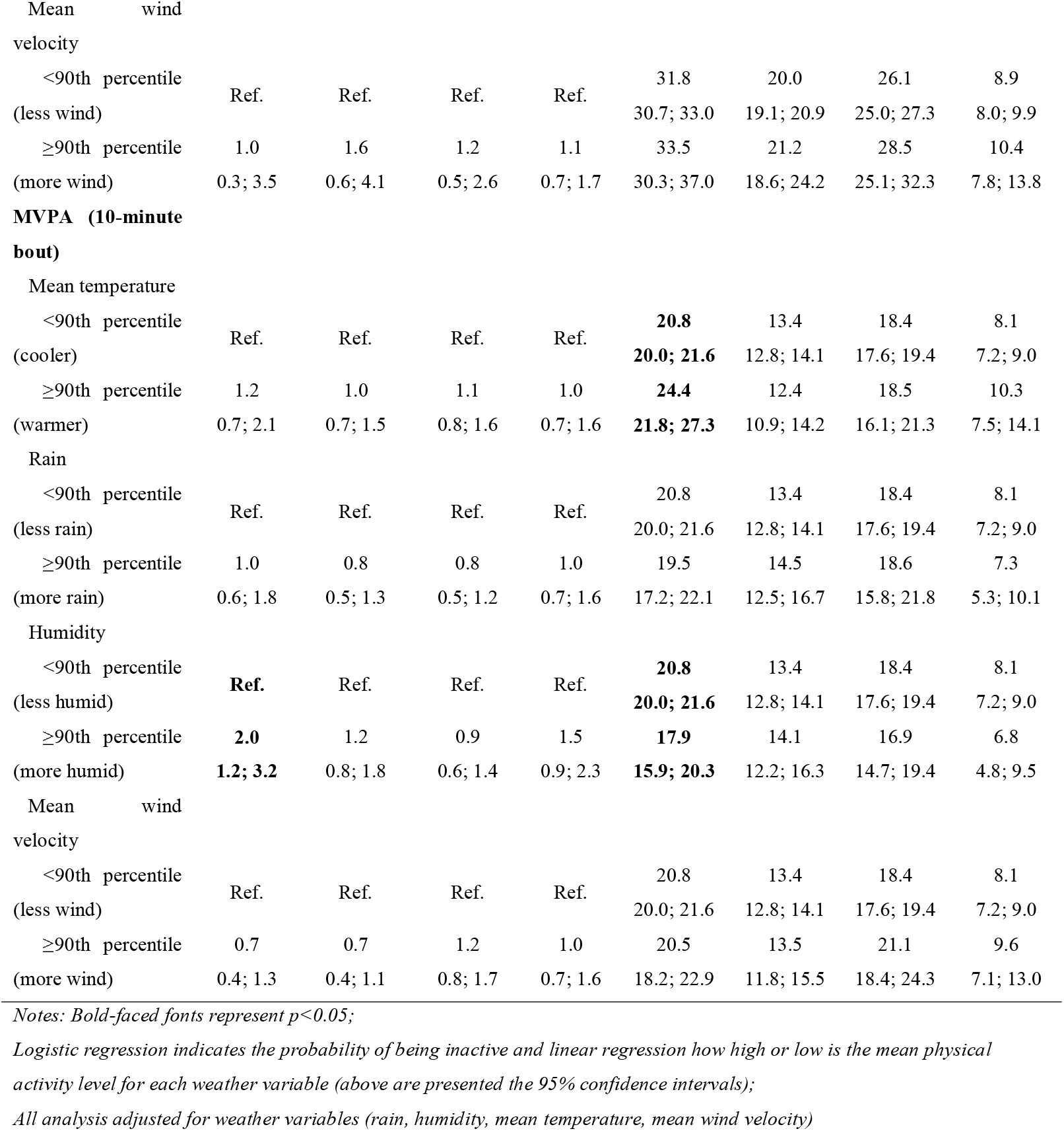
Adjusted association between weather variables and physical activity according to age estimated by two-part model

|  | Logistic Regression |  |  |  | Linear Regression |  |  |  |
| --- | --- | --- | --- | --- | --- | --- | --- | --- |
|  | Children | Young adults | Adults | Older adults | Children | Young adults | Adults | Older adults |
| **ENMO** |  |  |  |  |  |  |  |  |
| Mean temperature |  |  |  |  |  |  |  |  |
| <90th percentile | - | - | - | - | **65.3** | 33.4 | 35.3 | 21.6 |
| (cooler) | - | - | - | - | **64.6; 66.0** | 32.9; 33.9 | 34.8; 35.7 | 21.0; 22.2 |
| ≥90th percentile | - | - | - | - | **69.0** | 33.7 | 36.7 | 23.0 |
| (warmer) | - | - | - | - | **67.1; 70.9** | 32.3; 35.1 | 35.3; 38.0 | 21.3; 24.8 |
| Rain |  |  |  |  |  |  |  |  |
| <90th percentile | - | - | - | - | 65.3 | 35.3 | 64.2 | 35.3 |
| (less rain) | - | - | - | - | 64.6; 66.0 | 34.8; 35.7 | 62.0; 66.3 | 33.7; 36.8 |
| ≥90th percentile | - | - | - | - | 33.4 | 21.6 | 34.7 | 21.2 |
| (more rain) | - | - | - | - | 32.9; 33.9 | 21.0; 22.2 | 33.2; 36.3 | 19.5; 22.9 |
| Humidity |  |  |  |  |  |  |  |  |
| <90th percentile | - | - | - | - | **65.3** | 33.4 | 35.3 | 21.6 |
| (less humid) | - | - | - | - | **64.6; 66.0** | 32.9; 33.9 | 34.8; 35.7 | 21.0; 22.2 |
| ≥90th percentile | - | - | - | - | **61.0** | 32.7 | 35.2 | 20.7 |
| (more humid) | - | - | - | - | **58.9; 63.1** | 31.2; 34.1 | 33.9; 36.6 | 18.9; 22.4 |
| Mean wind velocity |  |  |  |  |  |  |  |  |
| <90th percentile | - | - | - | - | 65.3 | 33.4 | 35.3 | 21.6 |
| (less wind) | - | - | - | - | 64.6; 66.0 | 32.9; 33.9 | 34.8; 35.7 | 21.0; 22.2 |
| ≥90th percentile | - | - | - | - | 65.4 | 33.7 | 36.6 | 22.7 |
| (more wind) | - | - | - | - | 63.4; 67.3 | 32.2; 35.1 | 35.3; 38.0 | 21.1; 24.4 |
| **MVPA (5-minute bout)** |  |  |  |  |  |  |  |  |
| Mean temperature |  |  |  |  |  |  |  |  |
| <90th percentile | Ref. | Ref. | Ref. | Ref. | **31.8** | 20.0 | 26.1 | 8.9 |
| (cooler) |  |  |  |  | **30.7; 33.0** | 19.1; 20.9 | 25.0; 27.3 | 8.0; 9.9 |
| ≥90th percentile | 1.6 | 2.1 | 1.4 | 0.8 | **36.1** | 19.3 | 25.2 | 9.8 |
| (warmer) | 0.5; 4.7 | 1.0; 4.5 | 0.6; 2.9 | 0.5; 1.4 | **32.8; 39.8** | 17.1; 21.7 | 22.2; 28.6 | 7.3; 13.1 |
| Rain |  |  |  |  |  |  |  |  |
| <90th percentile | Ref. | Ref. | Ref. | Ref. | 31.8 | 20.0 | 26.1 | 8.9 |
| (less rain) |  |  |  |  | 30.7; 33.0 | 19.1; 20.9 | 25.0; 27.3 | 8.0; 9.9 |
| ≥90th percentile | 0.8 | 0.2 | 0.6 | 0.8 | 30.9 | 21.8 | 25.7 | 7.0 |
| (more rain) | 0.2; 3.0 | 0.0; 1.6 | 0.2; 2.0 | 0.5; 1.3 | 27.7; 34.4 | 19.1; 24.9 | 22.2; 29.8 | 5.3; 9.2 |
| Humidity |  |  |  |  |  |  |  |  |
| <90th percentile |  |  |  |  | **31.8** | 20.0 | 26.1 | 8.9 |
| (less humid) | Ref. | Ref. | Ref. | Ref. | **30.7; 33.0** | 19.1; 20.9 | 25.0; 27.3 | 8.0; 9.9 |
| ≥90th percentile | 2.1 | 1.3 | 0.5 | 1.0 | **25.9** | 18.4 | 24.6 | 6.7 |
| (more humid) | 0.7; 6.1 | 0.5; 3.8 | 0.2; 1.7 | 0.6; 1.6 | **23.3; 28.7** | 16.2; 20.9 | 21.7; 27.9 | 5.1; 9.0 |
| Mean wind velocity |  |  |  |  |  |  |  |  |
| <90th percentile (less wind) | Ref. | Ref. | Ref. | Ref. | 31.8<br>30.7; 33.0 | 20.0<br>19.1; 20.9 | 26.1<br>25.0; 27.3 | 8.9<br>8.0; 9.9 |
| ≥90th percentile (more wind) | 1.0<br>0.3; 3.5 | 1.6<br>0.6; 4.1 | 1.2<br>0.5; 2.6 | 1.1<br>0.7; 1.7 | 33.5<br>30.3; 37.0 | 21.2<br>18.6; 24.2 | 28.5<br>25.1; 32.3 | 10.4<br>7.8; 13.8 |
| **MVPA (10-minute bout)** |  |  |  |  |  |  |  |  |
| Mean temperature |  |  |  |  |  |  |  |  |
| <90th percentile (cooler) | Ref. | Ref. | Ref. | Ref. | **20.8**<br>**20.0; 21.6** | 13.4<br>12.8; 14.1 | 18.4<br>17.6; 19.4 | 8.1<br>7.2; 9.0 |
| ≥90th percentile (warmer) | 1.2<br>0.7; 2.1 | 1.0<br>0.7; 1.5 | 1.1<br>0.8; 1.6 | 1.0<br>0.7; 1.6 | **24.4**<br>**21.8; 27.3** | 12.4<br>10.9; 14.2 | 18.5<br>16.1; 21.3 | 10.3<br>7.5; 14.1 |
| Rain |  |  |  |  |  |  |  |  |
| <90th percentile (less rain) | Ref. | Ref. | Ref. | Ref. | 20.8<br>20.0; 21.6 | 13.4<br>12.8; 14.1 | 18.4<br>17.6; 19.4 | 8.1<br>7.2; 9.0 |
| ≥90th percentile (more rain) | 1.0<br>0.6; 1.8 | 0.8<br>0.5; 1.3 | 0.8<br>0.5; 1.2 | 1.0<br>0.7; 1.6 | 19.5<br>17.2; 22.1 | 14.5<br>12.5; 16.7 | 18.6<br>15.8; 21.8 | 7.3<br>5.3; 10.1 |
| Humidity |  |  |  |  |  |  |  |  |
| <90th percentile (less humid) | **Ref.** | Ref. | Ref. | Ref. | **20.8**<br>**20.0; 21.6** | 13.4<br>12.8; 14.1 | 18.4<br>17.6; 19.4 | 8.1<br>7.2; 9.0 |
| ≥90th percentile (more humid) | **2.0**<br>**1.2; 3.2** | 1.2<br>0.8; 1.8 | 0.9<br>0.6; 1.4 | 1.5<br>0.9; 2.3 | **17.9**<br>**15.9; 20.3** | 14.1<br>12.2; 16.3 | 16.9<br>14.7; 19.4 | 6.8<br>4.8; 9.5 |
| Mean wind velocity |  |  |  |  |  |  |  |  |
| <90th percentile (less wind) | Ref. | Ref. | Ref. | Ref. | 20.8<br>20.0; 21.6 | 13.4<br>12.8; 14.1 | 18.4<br>17.6; 19.4 | 8.1<br>7.2; 9.0 |
| ≥90th percentile (more wind) | 0.7<br>0.4; 1.3 | 0.7<br>0.4; 1.1 | 1.2<br>0.8; 1.7 | 1.0<br>0.7; 1.6 | 20.5<br>18.2; 22.9 | 13.5<br>11.8; 15.5 | 21.1<br>18.4; 24.3 | 9.6<br>7.1; 13.0 |

Notes: Bold-faced fonts represent $p < 0.05$ ;
Logistic regression indicates the probability of being inactive and linear regression how high or low is the mean physical activity level for each weather variable (above are presented the 95% confidence intervals);
All analysis adjusted for weather variables (rain, humidity, mean temperature, mean wind velocity)

## 4. Discussion

The present study described the association of weather variables on physical activity levels across different age groups (children, young adults, adults, and older adults). Physical activity levels were low, especially in older adults. Rain and mean wind velocity were not associated with physical activity of individuals, however, the probability of children being inactive was higher during humid days. After mutually adjusting the models for weather variables, only mean temperature and humidity were associated with higher and lower physical activity levels in children, respectively.

Our findings, in general, suggested that weather variables were not associated the probability of being physically inactive. Reasons for the lack of association include the complexity of shifting from a sedentary to an active status, since it comprises several aspects (e.g., individual, social, environmental, etc.), and not only weather factors.(6) The exception was for children, in which more humid days increased the probability of being inactive. It is important to highlight that the decision of being active or inactive in this age group differs from other groups – since they are more susceptible to parents’ and school’s decisions.(39,40) A series of studies have shown that the probability of being active is influenced by favorable environments (e.g. urban planning, transportation systems, and parks) and availability of public strategies and policies to encourage physical activity engagement at population level.(6,41,42) These determinants are even more relevant in middle-income countries, where opportunities are lower and inequalities are usually higher.(43,44)

In spite of non-significant results for young adults, adults and older adults, some variations in overall physical activity and MVPA levels according to weather variables were found. In general, there was a direct and indirect association of temperature and humidity with physical activity levels, respectively. The reduced physical activity in low temperature days could be explained by the lower convenience to engage in outdoor physical activities, and only individuals that exercise indoors remain active during such days.(45,46) While the positive association between physical activity levels and temperature is well describe among children,(47) adolescents(47) and adults,(48) the influence of humidity is less explored. Two studies found that humidity was associated to lower levels of vigorous activity among children and adolescents.(49,50) Authors highlighted that high humidity works as a possible modulator of temperature, generating a sensation of heat, exhaustion, and discomfort.(50)

In Pelotas, the Southern Brazilian city in which all four cohorts analyzed in this study were conducted, the relative humidity lies above 80% during most part of the year.(51) According to Köppen-Geiger’s climate classification, the city has a “Cfa” profile: rainy, with hot temperatures during the summer and high humidity in all seasons.(52) Hence, the marked seasons that are present in Pelotas might be a barrier for physical activity practice during the summer and winter. It was already described that days with milder weather features are more favorable to physical activity practice.(45) A strength of our study is that we were able to analyze weather variables on a daily basis, obtaining individual-level information about weather variables during the time that participants were wearing accelerometers. This allowed us to mutually adjust statistical models using different weather variables, evidencing that those weather variables do not account as one of the main predictors of physical activity, and the only weather factors that were relevant for physical activity practice in children were mean temperature and humidity.

Our study involves some limitations, including that physical activity was measured during a one-week period for each participant. Hence, if participants present a variable physical activity pattern throughout the year – depending on the weather pattern of the city –, the intra-individual variability would not be identified. A second limitation was the lack of information about the location (indoors or outdoors) in which physical activity was usually performed. This is relevant, as weather variables influence more outdoor activities than indoors. Also, the results should be interpretated in light of the sex difference among young adults from analyzed and excluded individuals.

Our study also presents some strengths, including the analysis of physical activity using two operationalizations – dichotomic (active or inactive) and continuous. This allowed us to better understand how weather variables influenced physical activity behavior while accounting for the increased number of zero minutes of physical activity. Regarding the exposures, we were able to obtain weather variables in a daily basis, a differential of our study from others that usually rely on season differences or more commonly used weather variables, such as temperature and rain. Also, we used four population-based cohorts, summing up an enormous analytic sample size with varying age groups. Lastly, a recent systematic review of school-aged children and adolescents (3-19 years) searched all the literature and included 26 studies. Only one of those were held in a middle-income country; the remaining 25 were from high income countries.(47) Although weather variables are not expected to change regarding the income level of the country, the physical activity patterns and availability of facilities might change. Thus, our study found that some weather variables were more important to certain age groups, but did not influence others. Finally, this study can help researchers to identify important weather variables to be added into their models, regardless of the physical activity operationalization they choose to use.

## Data Availability

All data produced in the present study are available upon reasonable request to the authors

