## Supplementary Table 1 for "Weather variation in objectively measured physical activity: results of four Brazilian cohorts"

Supplementary Table 1. Comparison between analyzed and excluded participants from each cohort.

|  | **Children** | | **Young adults** | | **Adults** | | **Elders** | |
| --- | --- | --- | --- | --- | --- | --- | --- | --- |
|  | **Analyzed**  **N (%)** | **Excluded**  **N (%)** | **Analyzed**  **N (%)** | **Excluded**  **N (%)** | **Analyzed**  **N (%)** | **Excluded**  **N (%)** | **Analyzed**  **N (%)** | **Excluded**  **N (%)** |
| Sex | *p=0.351* | | *p=0.003* | | *p=0.574* | | *p=0.205* | |
| Male | 1721 | 115 | 1397 | 386 | 1323 | 455 | 367 | 170 |
|  | (51.4) | (53.0) | (48.1) | (42.7) | (48.5) | (47.4) | (37.8) | (35.4) |
| Female | 1627 | 102 | 1509 | 518 | 1407 | 505 | 604 | 310 |
|  | (48.6) | (47.0) | (51.9) | (57.3) | (51.5) | (52.6) | (62.2) | (64.6) |
| Skin color | *p=0.011* | | *p<0.001* | | *p<0.001* | | *p=0.049* | |
| White | 2157 | 149 | 1726 | 536 | 2027 | 785 | 797 | 414 |
|  | (66.9) | (77.2) | (62.0) | (68.0) | (74.3) | (81.8) | (82.1) | (87.0) |
| Black | 414 | 16 | 460 | 78 | 460 | 114 | 126 | 42 |
|  | (12.8) | (8.3) | (16.5) | (9.9) | (16.9) | (11.9) | (13.0) | (8.8) |
| Other | 654 | 28 | 600 | 174 | 243 | 61 | 48 | 20 |
|  | (20.3) | (14.5) | (21.5) | (22.1) | (8.9) | (6.4) | (4.9) | (4.2) |
| Asset index | *p=0.231* | | *p=350* | | *p<0.001* | | *p=0.289* | |
| Q1 (poorest) | 651 | 54 | 605 | 180 | 686 | 181 | 227 | 105 |
|  | (19.7) | (25.2) | (20.9) | (20.2) | (26.3) | (20.2) | (24.3) | (23.1) |
| Q2 | 669 | 35 | 649 | 225 | 468 | 136 | 161 | 85 |
|  | (20.1) | (16.4) | (22.4) | (25.2) | (18.0) | (15.1) | (17.3) | (19.3) |
| Q3 | 660 | 47 | 525 | 146 | 528 | 171 | 169 | 78 |
|  | (20.0) | (21.9) | (18.2) | (16.4) | (20.3) | (19.0) | (18.2) | (17.7) |
| Q4 | 664 | 38 | 569 | 184 | 519 | 203 | 223 | 87 |
|  | (20.1) | (17.8) | (19.7) | (20.6) | (19.9) | (22.6) | (24.0) | (19.7) |
| Q5 (richest) | 665 | 40 | 542 | 157 | 405 | 207 | 151 | 86 |
|  | (20.1) | (18.7) | (18.8) | (17.6) | (15.5) | (22.1) | (16.2) | (19.5) |
